# Increased Prescribing of Psychotropic Medications to Ontario Nursing Home Residents during the COVID-19 Pandemic

**DOI:** 10.1101/2020.11.26.20239525

**Authors:** Nathan M. Stall, Jonathan S. Zipursky, Jagadish Rangrej, Aaron Jones, Andrew P. Costa, Michael P. Hillmer, Kevin Brown

## Abstract

In this population-based study of all Ontario nursing home residents, we found increased prescribing of psychotropic drugs at the onset of the COVID-19 pandemic that persisted through September 2020. Increases in prescribing were out of proportion to expected secular trends, and distinct from observed prescribing changes in other drugs during the pandemic. Our findings underscore the urgency of balancing infection prevention and control measures in nursing homes with the mental wellbeing of residents.

## Introduction

An enduring feature of the COVID-19 pandemic is the sizeable number of outbreaks and deaths in nursing homes. To limit the introduction of virus in these settings, strict and lengthy lockdowns were imposed on nursing homes worldwide. These restrictions have led to concerns about the mental health of nursing home residents, and increases in the use of psychotropic medications, which were frequently overprescribed prior to the pandemic.^1,2^

## Methods

We examined the monthly proportion of nursing home residents in Canada’s most populous province of Ontario dispensed psychotropics from January–September 2020. As of March 1, 2020, there were 77,291 residents living in the province’s 623 licensed nursing homes receiving personal and nursing care, subsidized accommodation and all prescription medications under a publicly long-term care program. Restrictions on visitation, absences, and most congregate dining and group activities began in all nursing homes on March 14, 2020.

We studied the dispensation of four psychotropic medications: antipsychotics (1^st^-and-2^nd^ generation), antidepressants (SSRIs, SNRIs, bupropion, mirtazapine), benzodiazepines, and trazodone. We also examined the prescribing of two negative tracer drugs (metformin and statins) not expected to change during the pandemic. Data were obtained from the Ontario Drug Benefit database through the Ministry of Health as part of the province’s COVID-19 Modeling Consensus Table.

We examined the proportion of residents receiving a prescription across the first seven months of the pandemic (March-September 2020). To account for the dynamic number of residents living in nursing homes during the pandemic, we calculated the monthly number of residents filling a prescription for each drug of interest as a proportion of the monthly number of residents filling a prescription for any drug. In unadjusted analyses, we examined absolute changes in the proportion of residents receiving a prescription compared to the most recent pre-pandemic months (January-February 2020). In adjusted analyses, we examined absolute changes compared to anticipated absolute prescribing trends based on the linear secular trend of prescribing rates from April 2019 to February 2020. All analyses were based on logistic regression. The study was approved by the University of Toronto’s research ethics board.

## Results

Comparing January-February 2020 with September 2020 (Table 1), there were absolute increases in the proportion of nursing home residents who received prescriptions for antipsychotics (1.6%, 95% Confidence Internal (CI): 0.9–2.4), antidepressants (1.6%, 95%CI: 1.0–2.2), and trazodone (1.0%, 95%CI: 0.3–1.8). While benzodiazepine prescribing decreased slightly (−0.2%, 95%CI: - 1.0–0.6), the adjusted analysis (Figure 1) revealed an increase relative to apparent negative secular trends (0.3%, 95%CI: −0.3–0.9). Deviations above positive secular trends were also present in adjusted analyses for all other psychotropics, including antipsychotics (1.7%, 95%CI: 1.1–2.2), antidepressants (1.4%, 95%CI: 0.9–1.8), and trazodone (1.1%, 95%CI: 0.6–1.7). Comparatively, there were decreases in the proportion of residents prescribed metformin (−0.3%, 95%CI: −1.1–0.5) and statins (−0.7%, 95%CI: −1.4–0.1), and these decreases were more pronounced after adjustment for secular trends (−0.6%, 95%CI:-1.2–0.0 and −1.4%, 95%CI: −1.9– −0.8, respectively).

**Table 1:** Unadjusted and adjusted absolute changes in psychotropic drug prescribing to nursing home residents in Ontario, Canada (January-September, 2020)

|  | Jan-Feb, 2020 | Mar-Sep, 2020 |  |  | Sept, 2020 |  |  |
| --- | --- | --- | --- | --- | --- | --- | --- |
|  | Mean monthly proportion<sup>1</sup> of residents receiving prescription (%) | Mean monthly proportion of residents receiving prescription (%) | Unadjusted difference, % (95% CI)<sup>2</sup> | Adjusted difference, % (95% CI)<sup>3</sup> | Monthly proportion<sup>1</sup> of residents receiving prescription (%) | Unadjusted difference, % (95% CI)<sup>2</sup> | Adjusted difference, % (95% CI)<sup>3</sup> |
| **Psychotropics** |  |  |  |  |  |  |  |
| Antidepressants | 51.3 | 52.3 | 1.0 (0.6, 1.4) | 0.8 (0.5, 1.1) | 52.9 | 1.6 (1.0, 2.2) | 1.4 (0.9, 1.8) |
| Trazodone | 25.4 | 26.1 | 0.7 (0.2, 1.2) | 0.7 (0.3, 1.0) | 26.5 | 1.0 (0.3, 1.8) | 1.1 (0.6, 1.7) |
| Antipsychotics | 28.4 | 29.5 | 1.1 (0.6, 1.6) | 1.0 (0.7, 1.3) | 30.0 | 1.6 (0.9, 2.4) | 1.7 (1.1, 2.2) |
| Benzodiazepines | 11.3 | 11.2 | -0.1 (-0.6, 0.5) | 0.2 (-0.2, 0.5) | 11.1 | -0.2 (-1.0, 0.6) | 0.3 (-0.3, 0.9) |
| **Negative tracers** |  |  |  |  |  |  |  |
| Metformin | 12.7 | 12.5 | -0.2 (-0.7, 0.3) | -0.3 (-0.7, 0.0) | 12.4 | -0.3 (-1.1, 0.5) | -0.6 (-1.2, 0.0) |
| Statins | 31.0 | 30.3 | -0.7 (-1.2, -0.2) | -0.8 (-1.1, -0.5) | 30.3 | -0.7 (-1.4, 0.1) | -1.4 (-1.9, -0.8) |

<sup>1</sup> The monthly number of residents filling a prescription of interest as a proportion of the monthly number of residents filling a prescription for any drug. As of March 1, 2020 there were 77,291 residents living in Ontario's 623 licensed nursing homes.
<sup>2</sup> Unadjusted absolute difference with 95% Confidence Interval (CI) compared to January-February, 2020.
<sup>3</sup> Adjusted absolute difference with 95% Confidence Interval (CI) compared to prescribing rates expected based on a linear secular trend in the April 2019 to February 2020 period.

**Figure 1:**
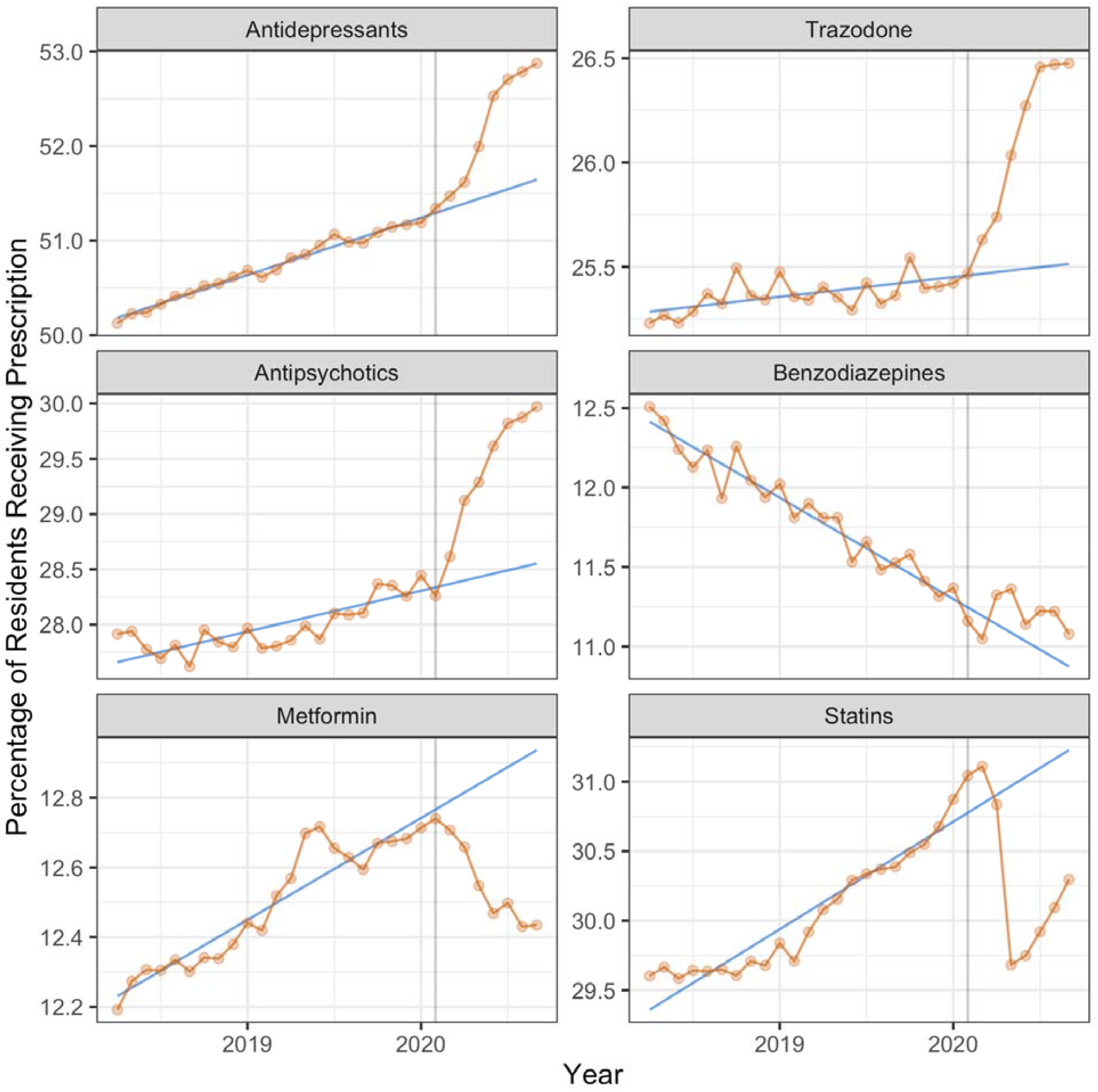
Changes in psychotropic drug prescribing to nursing home residents in Ontario, Canada (April 2019 through September 2020) Monthly proportion of residents filling a prescription for each drug class from April 2019 to September 2020. As of March 1, 2020, there were 77,291 residents living in Ontario’s 623 licensed nursing homes. The grey vertical line represents the onset of the COVID-19 lockdown in Ontario (March 2020). The blue line represents expected prescribing based on secular trends prior to the lockdown.

## Discussion

In this population-based study of all Ontario nursing home residents, we found increased prescribing of psychotropic drugs at the onset of the COVID-19 pandemic that persisted through September 2020. Increases in prescribing were out of proportion to expected secular trends, and distinct from observed prescribing changes in other drugs during the pandemic.

Our results align with findings of increased prescribing of antipsychotics during the pandemic to people living with dementia in the United Kingdom.^3^ We offer additional insights by studying prescribing of multiple classes of psychotropic drugs, as well as those unexpected to change during the pandemic. Our study is limited by lack of data on prescribing indications; while some of the observed increases in psychotropics may be for palliative management of residents dying of COVID-19, increases were more likely related to the effects of prolonged social isolation during lockdowns.^4,5^ Due to diminished resources, clinicians may have been less likely to prioritize more favorable non-pharmacological management for common issues like responsive behaviours. Overall, our findings underscore the urgency of balancing infection prevention and control measures in nursing homes with the mental wellbeing of residents.^6^

## Data Availability

Study protocol and statistical code: The study protocol and underlying analytic code are available from the authors on request (e-mail), with the understanding that the computer programs may rely on coding templates or macros that are unique to the Ontario Ministry of Health and therefore either are inaccessible or may require modification. Data set: The data used for this study are not publicly available.

